# Urine γ-interferon-inducible protein (IP-10) as a biomarker of histological activity of lupus nephritis

**DOI:** 10.1101/2022.08.16.22278832

**Authors:** Karan Prasopsanti, Thanarat Supasiri, Yingyos Avihingsanon, Kroonpong Iampenkhae, Jerasit Surintrspanont, Yuda Chongpisan, Chutipha Promjean, Thitima Benjachat Suttichet, Theerada Assawasaksakul, Nont Oudomying, Wonngarm Kittanamongkolchai

**Affiliations:** Division of Rheumatology, Department of Medicine, Faculty of Medicine, Chulalongkorn University, Bangkok, Thailand; Queen Savang Vadhana Memorial Hospital; Division of Nephrology, Department of Medicine, Faculty of Medicine, Chulalongkorn University, Bangkok, Thailand; Renal Immunology and Therapeutic Apheresis Research Unit, Faculty of Medicine, Chulalongkorn University, Bangkok, Thailand; Department of Pathology, Faculty of Medicine, Chulalongkorn University, Bangkok, Thailand; Biostatistics Excellence Center, Research Affairs, Faculty of Medicine, Chulalongkorn University; Maha Chakri Sirindhorn Clinical Research Center, Faculty of Medicine, Chulalongkorn University, Bangkok, Thailand; Clinical Pharmacokinetics and Pharmacogenomics Research Unit, Faculty of Medicine, Chulalongkorn University; Chulalongkorn University International Medical Program (CU-MEDi), Faculty of Medicine, Chulalongkorn University, Bangkok, Thailand

**Keywords:** Lupus nephritis, biomarker, IP-10, gamma-interferon

## Abstract

**Introduction:** Conventional markers are not reliable predictors of histological activity of lupus nephritis (LN). We aimed to examine the utility of urine γ-interferon-inducible protein (IP-10) in predicting LN flares, diagnosis of LN, and forecasting treatment response.

**Methods:** SLE patients who fulfilled the ACR 1997 criteria with history of LN were enrolled. Urine IP-10 was measured at least once during routine quarterly visits, at the time of diagnosis of active LN, and monthly during induction therapy for 6 months.

**Results:** There were 65 active LN and 46 inactive LN included. The mean urine IP-10 levels among the active and inactive LN were 2.69 (95%CI 2.53-2.86) and 2.18 (95%CI 1.96-2.39) log copies/mcg total RNA respectively (p-value < 0.0001). Clinicopathological discordance was observed in 9 of 55 (16%) biopsied patients (5 with proliferative LN without proteinuric flare and 4 with nephrotic-range proteinuria from glomerulosclerosis). Urine IP-10 predicted histological activity of LN with 91% accuracy, compared to 84% with proteinuric flare. Within two years, half of the clinically inactive LN patients with positive baseline urine IP-10 developed LN flare, whereas no flares were observed in patients with negative baseline urine IP-10. Urine IP-10 levels were not associated with treatment response at 6 months.

**Conclusion:** Urine IP-10 may reflect histological activity of LN more accurately than conventional markers, especially in patients with clinicopathologic discrepancy. Clinically inactive LN patients with positive urine IP-10 were at a higher risk of developing LN flare.

**Key messages:**

- The majority of the studies on novel biomarkers in LN lacked renal biopsy and relied on clinical indicators to determine histological activity. As a result, the validity of these studies may be jeopardized.
- According to this study, clinicopathological discordance was found in 16% of LN patients who underwent renal biopsy. Urine IP-10 outperformed urinary protein level in differentiating histologically active LN from inactive LN (accuracy 91% versus 84%).
- Within two years, half of the clinically inactive LN patients who had positive urine IP-10 developed LN flares, whereas none of those who had negative urine IP-10 did.
- Urine IP-10 may aid in the diagnosis of histologically active LN, particularly in patients with clinicopathologic discrepancy. Urine IP-10 monitoring in clinically inactive LN patients may predict the risk of future LN flares.

## Introduction

Systemic lupus erythematosus (SLE) is a complex autoimmune disease characterized by unpredictable flares of disease activity and irreversible damage to multiple organ systems. Despite the availability of aggressive immunosuppressive therapies, up to half of patients with aggressive lupus nephritis (LN) develop end-stage kidney disease within 15 years.(1) Early detection of active LN with prompt initiation of immunosuppressive therapy is the key to improve renal outcome and survival rate.(2, 3) Unfortunately, determining renal disease activity in LN remains a challenge given that current biomarkers are neither sensitive nor specific.(4) Although proteinuria is the main criteria for LN flare, it can be a result of chronic renal damage and not necessarily an indication of ongoing inflammation in the kidneys. On the other hand, studies have shown that LN occurs in one third of SLE patients without a significant increase in proteinuria.(5) Although renal biopsy remains the gold standard, it is invasive and cannot be performed repeatedly. All in all, identification of novel biomarkers that reflect renal disease activity remains an unmet need.

γ-interferon-inducible protein (IP-10), T-helper 1 type chemokines, also known as C-X-C motif chemokine ligand 10 (CXCL10), is one of the potential novel biomarkers for active LN. It plays a critical role in the infiltration of inflammatory cells into the affected organs in SLE patients.(6-8) In a lupus mouse model, this chemokine regulates the Th1-cell migration into the kidneys and lungs via interaction with its corresponding chemokine receptor, chemokine (C-X-C motif) receptor 3 (CXCR3).(7) Neutralizing monoclonal antibodies or small-molecule inhibitors that disrupts CXCR3’s function markedly attenuates the inflammatory response, thereby reducing kidney damage.(9)

Our previous study in 26 patients with LN showed that urine IP-10 could distinguish severe proliferative forms of LN among others, with an accuracy greater than the current available clinical markers. In addition, Urine IP-10 was also found to be higher before a LN flare and then declined in response to treatments.(10) While some prior studies showed consistent findings,(11, 12, 13) others reported no difference in urine IP-10 levels between active and inactive LN.(13, 14) Several factors including a small number of patients, the absence of pathological confirmation of LN and the use of cross-sectional design may have contributed to these conflicting results.

In this study, we aimed to validate the utility of urine IP-10 in flare prediction, diagnosis of LN and treatment response prediction among biopsy-proven LN patients with a longitudinal follow-up.

## Materials and Methods

### Patients

The study was approved by the ethics committee of the Faculty of Medicine, Chulalongkorn University. Patients with a diagnosis of SLE, fulfilling American College of Rheumatology (ACR) 1997 criteria with history of biopsy-proven Class III, IV or V LN, were recruited from the specialized lupus nephritis clinic, King Chulalongkorn Memorial Hospital, Bangkok, Thailand from 1^st^ January 2013 to 31^st^ December 2018. Exclusion criteria included inadequate clinical data (lack of serum creatinine or urinary protein/creatinine index (UPCI)), pregnancy, HIV infection, end stage renal disease, and poor quality of urine sample (abnormally cloudy urine or samples with 18s rRNA less than 1000 copies per microgram of total RNA). Nine healthy volunteers were recruited as controls.

### Classification of SLE activity status

Patients with active LN had either 1) histologically active LN, defined as biopsy-proven LN class III, IV, or V according to the ISN/RPS 2003 classification of LN, or 2) clinically active LN if a renal biopsy was unavailable, defined as an increase in serum creatinine of more than 25% after excluding other potential causes, and/or experiencing a proteinuric flare (an increase in urinary protein/creatinine index (UPCI) to greater than 1 g/g if baseline UPCI < 0.5 g/g, or an increase of ≥ 2 folds if baseline UPCI ≥ 0.5 g/g).

Patients with inactive LN were defined as having either 1) histologically inactive LN (no proliferative or membranous lesions on biopsy), or 2) clinically inactive LN (did not meet clinically active LN criteria if renal biopsy was not performed).

Treatment response was defined by normal or ≤25% decline of eGFR(Chronic Kidney Disease Epidemiology Collaboration equation) from baseline and at least a 50% reduction of proteinuria, with a UPCI between 0.5 to 3 g/g.

### Follow-up schedule and treatment regimen

Active LN patients were followed up monthly for 6 months while receiving induction therapy and quarterly thereafter. Every visit, the complete blood count, serum albumin, blood urea nitrogen, serum creatinine, UPCI, and urinalysis were measured. Urine IP-10 measurements were performed at least once during routine quarterly visits, at the time of diagnosis of active LN, and monthly during induction therapy for 6 months.

Standard induction therapy was either cyclophosphamide (National Institute of Health or Euro-Lupus regimen) or mycophenolate mofetil (1.5-2 g/day). Oral prednisolone was initiated at 0.5-1 mg/kg/day then reduced by 5 mg/day after the second and fourth week, then reduced again by 5 mg/day every 4 weeks until a dosage of 5 mg/day was reached. Standard maintenance therapy was either azathioprine (1-2 mg/kg/day) or mycophenolate mofetil (1-1.5 g/day).

### Urinary-cell level of IP-10 mRNA measurement

Fresh urine samples were immediately centrifuged at 1000 x g for 30 minutes at 4 degrees Celsius. Total RNA was extracted from urinary cell pellets using an RNA blood mini kit (Qiagen, Valencia, CA, USA). All extracted urine RNAs have undergone quality and quantity checking by using NanoDrop™ Spectrophotometers (Thermo Fisher Scientific Inc., Waltham, MA, USA). Ratio of absorbance at 260 nm to 280 nm was used to measure the purity of RNA extractions, where 2.0 was considered to be the best quality which could be used for subsequent analysis. Two hundred and fifty nanograms of total RNA were reverse-transcribed into complementary DNA (cDNA) by using MultiScribe™ Reverse Transcriptase enzyme (Applied Biosystems, Carlsbad, CA, USA). The urinary IP-10 mRNA level was quantified by a real-time PCR technique (Light Cycler, Roche Molecular Biochemicals, Indianapolis, IN, USA). The housekeeping gene, 18s rRNA, was measured as a reference gene.(10) Samples that had 18s rRNA less than 1000 copies per microgram of total RNA indicated low quality and were excluded.

### Statistical analyses

Quantitative variables with normal distribution were expressed as mean (95% CI) unless otherwise indicated. Comparisons of the baseline characteristics and clinical parameters between active LN and inactive LN were evaluated using chi-square and independent samples t-test. Urine IP-10 levels for both active and inactive LN were compared with healthy controls using one-way ANOVA. Associations of urine IP-10 and clinical parameters were tested using a linear regression model. Univariable and multivariable logistic regression were performed to determine the association of urine IP-10 and other conventional markers with LN activity. Only variables with p <0.05 after performing univariable analysis were included in the multivariable analysis. Receiver operating characteristic (ROC) plots were used to evaluate the performance of urine IP-10 in predicting active LN. Sensitivity and specificity derived from the ROC curves were used to identify the cut-off point for urine IP-10 level with Youden’s index criteria. The optimal cut-off point determined from ROC analysis defined a positive IP-10. A p-value less than 0.05 was considered as statistical significance. Statistical analysis was performed using JMP software (SAS Institute, Cary, North Carolina, USA) and dot-plot graphs were created using GraphPad Prism V.4.03 (GraphPad Software, La Jolla, CA, USA).

## Results

Of 178 SLE patients with a history of LN seen in the specialized LN clinic, 145 provided urine samples for IP-10 measurement. Thirty-four patients were excluded due to insufficient clinical data, pregnancy, HIV infection, ESRD, or poor quality urine samples (**Figure 1**). There were 111 patients included in the study, 65 of whom had active LN (51 with histologically active LN) and 46 patients with inactive LN (4 with histologically inactive LN). Thirty-six patients with active LN were followed monthly for six months during induction therapy. Fifteen clinically inactive patients were followed for up to two years. **Table 1** displays the demographic characteristics of the patients.

**Table 1.** Demographic characteristics of active and inactive lupus nephritis patients

| Clinical parameters | Active lupus nephritis<br>(N=65) | Inactive lupus<br>nephritis (N=46) | p-value |
| --- | --- | --- | --- |
| Age (years) | 34.3 (10.8) | 36.3 (11.4) | 0.4 |
| Sex (M:F) | 5:60 | 3:43 | 0.8 |
| Immunosuppressive treatment at the time of<br>urine IP-10 measurement |  |  | 0.5 |
| - Mycophenolate mofetil | 24 (37%) | 12 (26%) |  |
| - Azathioprine | 9 (14%) | 7 (15%) |  |
| - Prednisolone alone | 32 (49%) | 27 (59%) |  |
| Mean dosage of immunosuppressive<br>treatment at the time of urine IP-10<br>measurement (mg/day) |  |  |  |
| - Mycophenolate mofetil | 1219 (557) | 942 (435) | 0.1 |
| - Azathioprine | 56 (17) | 46 (10) | 0.2 |
| - Prednisolone | 16 (16) | 7 (8) | 0.0004 |
| Hemoglobin (g/dL) | 10.8 (1.8) | 11.8 (1.6) | 0.003 |
| White blood cell (10*3/ul) | 6,534 (3,528) | 6,842 (2,705) | 0.6 |
| Platelet (10*3/ul) | 245,543 (82,458) | 243,050 (77,435) | 0.9 |
| Serum creatinine (mg/dL) | 1.18 (0.72) | 0.87 (0.28) | 0.002 |
| Serum albumin (g/dL) | 2.97 (0.60) | 3.89 (0.51) | <0.000<br>1 |
| UPCI (g/g Creatinine) | 4.24 (5.54) | 0.77 (1.00) | <0.000<br>1 |
| Urine red blood cell (cell/HPF) | 13.5 (18.1) | 4.9 (14.6) | 0.007 |
| Urine white blood cell (cell/HPF) | 12.9 (12.3) | 7.3 (16.5) | 0.06 |
| C3 (mg/dL) (median, IQR) | 40 (36-70) | 83 (68-92) | 0.004 |
| C4 (mg/dL) (median, IQR) | 6 (3-20) | 18 (9-29) | 0.1 |
| Anti-dsDNA (IU/ml) (median, IQR) | 287 (112-783) | 115 (4-394) | 0.2 |

Data expressed as mean (standard deviation) unless otherwise stated, UPCI: urine protein-creatinine index, HPF: high power field, IQR: interquartile range

**Figure 1:**
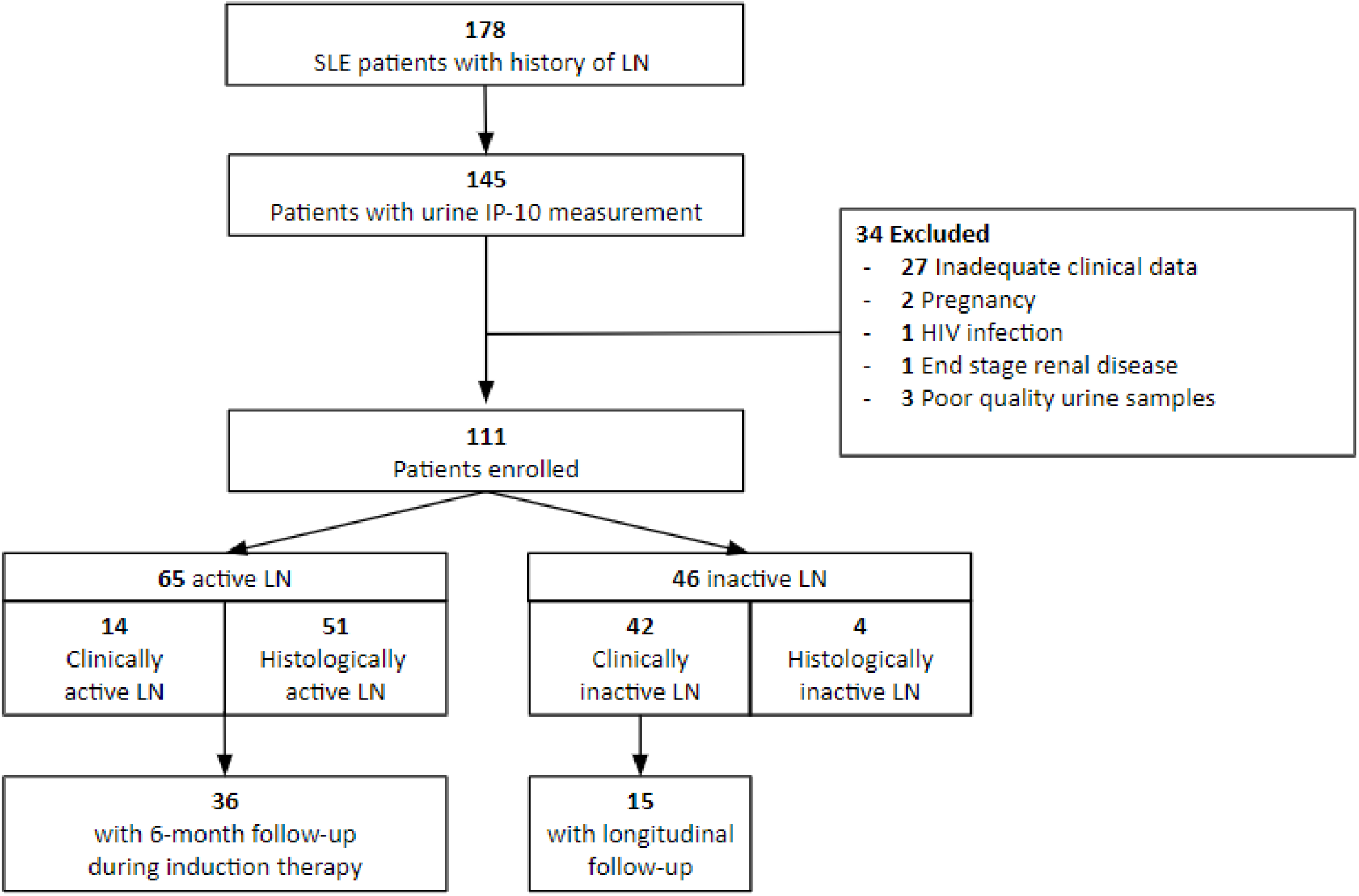
Flow diagram - Active LN included patients with histologically active LN or clinically active LN if a renal biopsy was unavailable - Histologically active LN defined as biopsy-proven LN class III, IV, or V according to the ISN/RPS 2003 classification of LN - Clinically active LN defined as an increase in serum creatinine of more than 25% after excluding other potential causes, and/or experiencing a proteinuric flare (an increase in UPCI to greater than 1 g/g if baseline UPCI < 0.5 g/g, or an increase of ≥ 2 folds if baseline UPCI ≥ 0.5 g/g). - Inactive LN included patients with either histologically inactive or clinically inactive LN - Histologically inactive LN defined as no proliferative or membranous lesions on biopsy - Clinically inactive LN included patients who did not meet clinically active LN criteria

### The relationships between urine IP-10 and clinical parameters

Urine IP-10 levels were weakly correlated with white blood cells (R^2^ =0.04, p-value 0.03) and urine white blood cells (R^2^ =0.06, p-value 0.008). There were no relationships found between urine IP-10 level and hemoglobin, platelet, serum creatinine, serum albumin, urine red blood cells, UPCI, C3, C4, or Anti-dsDNA.

### Predictive performance of urine IP-10 to differentiate active LN

The mean urine IP-10 was highest in active LN patients (2.69 log copies/mcg total RNA, 95% CI 2.53-2.86), followed by inactive LN (2.18 log copies/mcg total RNA, 95%CI 1.96-2.39) and lowest in healthy controls (0.89 log copies/mcg total RNA, 95% CI 0.22-1.57) (p-value < 0.0001) (**Figure 2**). Results from the univariable analysis showed that the odds(OR) of active LN increased by 3.03 (95% CI 1.61-5.67, p=0.0006) times for every unit of increasing urine IP-10 (**Table 2**). When adjusted for other variables included in the multivariable analysis, the OR for urine IP-10 rose over 5-fold to 16.64 (95% CI 3.58-77.3, p=0.0003).

**Table 2.** Univariable and multivariable analysis of clinical and laboratory parameters’ association with LN activity

| Variable | Univariable analysis |  |  | Multivariable analysis |  |  |
| --- | --- | --- | --- | --- | --- | --- |
|  | Odds ratio | 95% CI | p-value | Odds ratio | 95% CI | p-value |
| Age(year) | 0.97 | 0.95-1.02 | 0.4 |  |  |  |
| Hemoglobin(g/dL) | 0.70 | 0.55-0.90 | 0.005 | 1.67 | 0.78-3.60 | 0.2 |
| White blood cell (10*3/ul) | 0.99997 | 0.9998-1.00 | 0.6 |  |  |  |
| Platelet (10*3/ul) | 1.00 | 0.9999-1.00 | 0.9 |  |  |  |
| Urine IP-10 (log copies/mcg total RNA) | 3.03 | 1.61-5.70 | 0.0006 | 16.64 | 3.58-77.31 | 0.0003 |
| UPCI (g/g) | 3.12 | 1.90-5.14 | <0.0001 | 3.21 | 1.35-7.61 | 0.008 |
| Serum creatinine (mg/dL) | 4.05 | 1.40-11.79 | 0.01 | 8.68 | 1.15-65.70 | 0.04 |
| Serum albumin (g/dL) | 0.03 | 0.01-0.12 | <0.0001 | 0.01 | 0.0006-0.18 | 0.002 |
| Urine red blood cell (per 10 cells/hpf) | 1.06 | 1.01-1.11 | 0.02 | 1.06 | 1.00-1.12 | 0.049 |
| Urine white blood cell (per 10 cells/hpf) | 1.04 | 0.99-1.08 | 0.06 |  |  |  |

UPCI: urine protein-creatinine index, HPF: high power field

**Figure 2.**
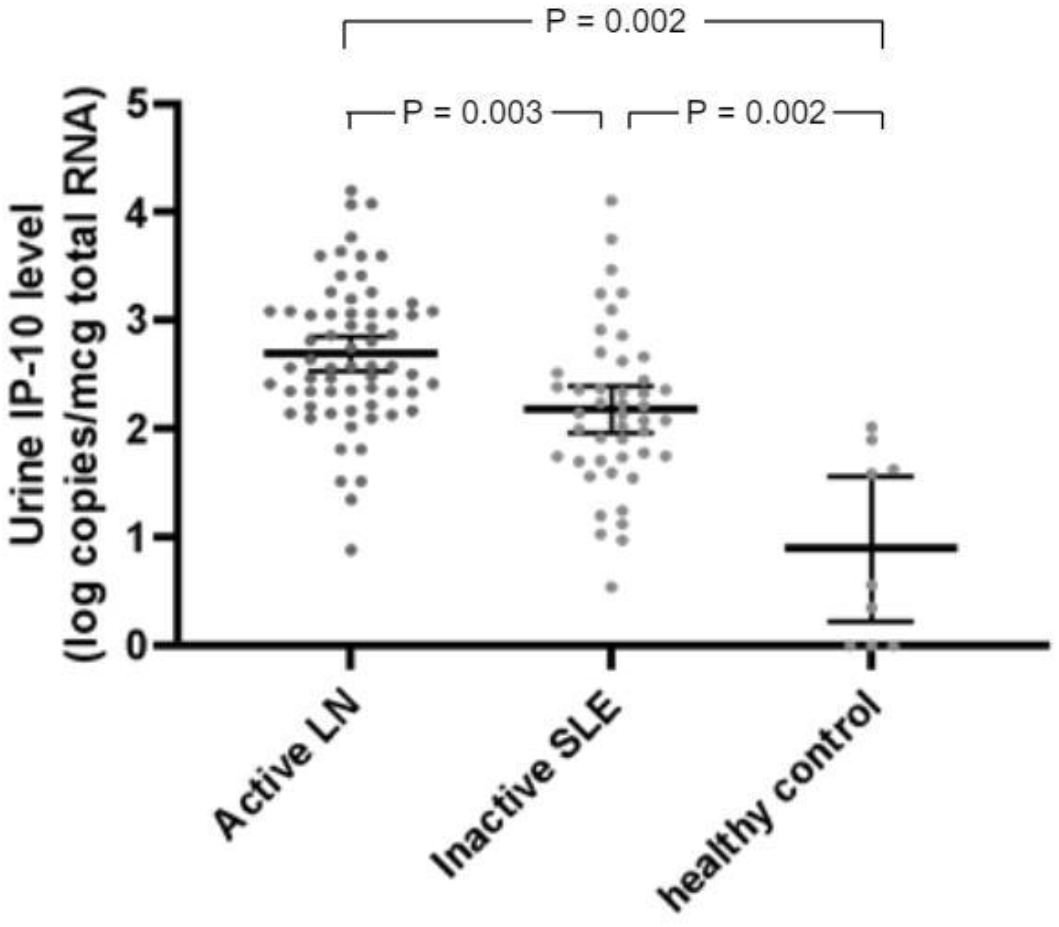
Urine IP-10 levels in patients with active lupus nephritis, inactive lupus nephritis and healthy controls.

ROC curve analysis was used to assess the diagnostic accuracy of urine IP-10. The ROC-AUC of urine IP-10 in predicting active LN was 0.71 (95% CI 0.61-0.81, p=0.0002). The urine IP-10 level of 2.10 log copies/mcg total RNA yielded the highest overall sensitivity of 89% and specificity of 43%, and was selected as a cut-off value.

### The role of urine IP-10 in LN with clinicopathological discrepancy

A renal biopsy was performed on 55 of the 111 patients. Fifty-one patients had histologically active LN (LN Class III = 15, Class IV = 26, Class III+V = 6, and Class IV +V = 4), while four had histologically inactive LN. Nine patients (16%) had clinicopathological discordance, with 5 having histologically active LN without proteinuric flare (an increase in UPCI to greater than 1 g/g, if baseline UPCI < 0.5 g/g, or an increase of ≥ 2 folds, if baseline UPCI ≥ 0.5 g/g) and 4 having histologically inactive LN with proteinuric flare.

Urine IP-10 accurately predicted histological activity in 50 (91%) patients using a cut-off value of 2.10 log copies/mcg total RNA, compared to 46 patients (84%) when using proteinuric flare(**Table 3**). The majority of patients with histologically active LN had both positive urine IP-10 and proteinuric flare (N = 43, 84%), while 4 (8%) had only positive urine IP-10, 3 (6%) had only proteinuric flare, and 1 (0.02%) had neither. Urine IP-10 levels were negative in three out of four patients with histologically inactive LN and nephrotic range proteinuria (**Table 3**).

**Table 3.** Contingency table comparing urine IP-10 and proteinuric flare in patients with a) histologically active LN and b) histologically inactive LN

**a) Histologically active LN**
|  | Positive<br>urine IP-10 | Negative<br>urine IP-10 | Total |
| --- | --- | --- | --- |
| With proteinuric<br>flare | 43 | 3 | 46 |
| Without<br>proteinuric flare | 4 | 1 | 5 |
| Total | 47 | 4 | 51 |

**b) Histologically inactive LN**
|  | Positive<br>urine IP-10 | Negative<br>urine IP-10 | Total |
| --- | --- | --- | --- |
| With proteinuric<br>flare | 1 | 3 | 4 |
| Without<br>proteinuric flare | 0 | 0 | 0 |
| Total | 1 | 3 | 4 |

- Cut-off value of urine IP-10 = 2.10 log copies/mcg total RNA - Proteinuric flare defined as an increase in UPCI to greater than 1 g/g if baseline UPCI < 0.5 g/g, or an increase of $\geq 2$ folds if baseline UPCI $\geq 0.5$ g/g - Histologically active LN defined as biopsy-proven LN class III, IV, or V according to the ISN/RPS 2003 classification of LN - Histologically inactive LN defined as no proliferative or membranous lesions on renal biopsy

The clinical characteristics of histologically active LN patients with only positive urine IP-10 or only proteinuric flare are shown in **Table 4**. Patients with only positive urine IP-10 without proteinuric flare had preserved podocyte structure despite coexisting severe renal inflammation. Most of them were given low-dose immunosuppressive therapy, and their renin-angiotensin-converting enzyme inhibitor dosage was stable prior to the renal biopsy. After induction therapy, all patients had stable renal function and normalization of serum complement without clinical LN flare at the 12-month follow-up. On the other hand, patients with proteinuric flare with negative urine IP-10 had diffused podocyte foot process effacement. This group of patients received higher doses of immunosuppressive treatment at the time of urine IP-10 measurement when compared to those with only positive urine IP-10 (median dose of prednisolone 55 (IQR 50-60) mg/day versus 10 (IQR 3-26) mg/day).

**Table 4.**
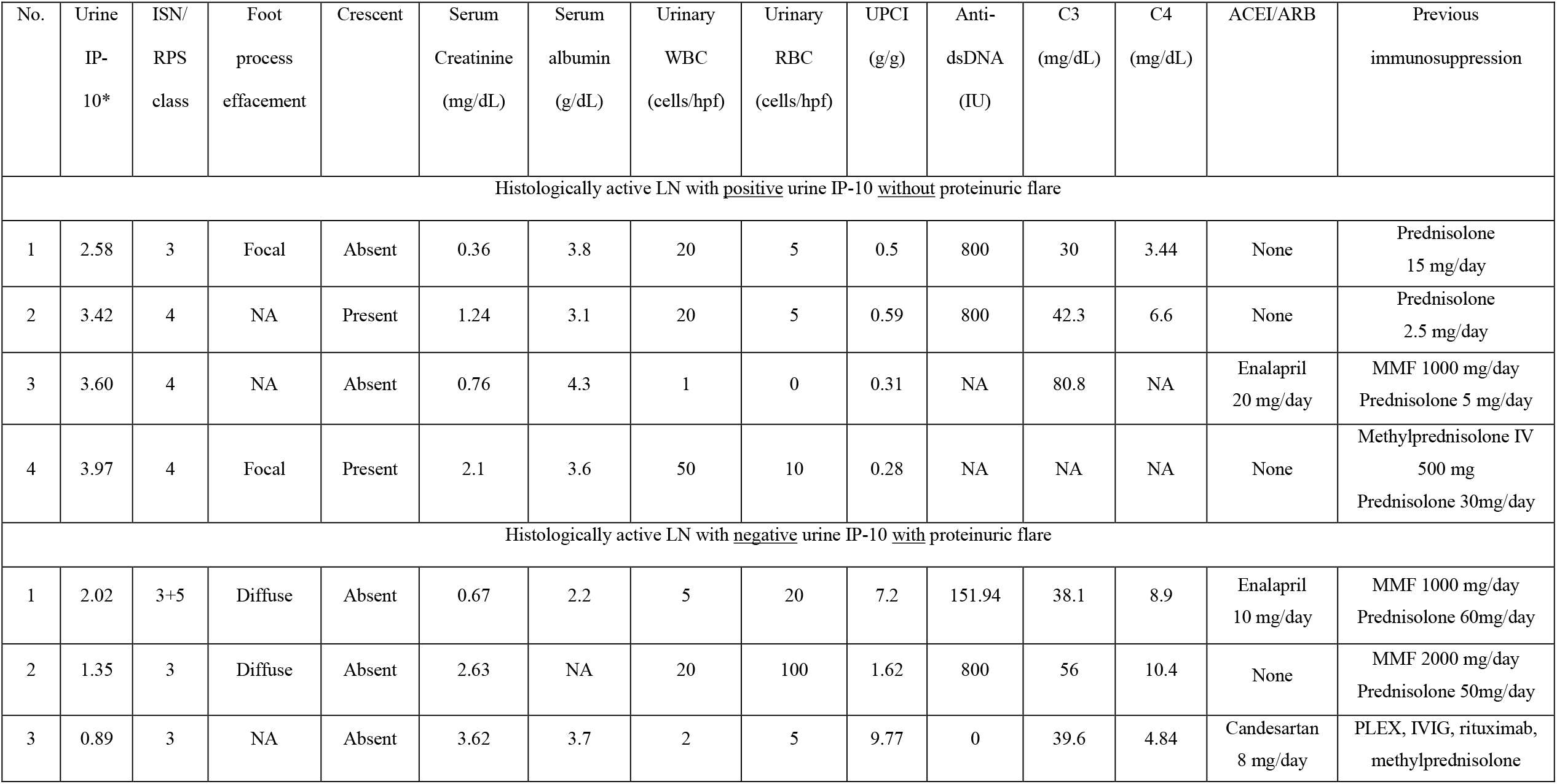
The clinical characteristics of patients with histologically active LN with only positive urine IP-10 or only proteinuric flare ISN/RPS: International Society of Nephrology/Renal Pathology Society 2003 classification of lupus nephritis and 2018 revision, UPCI: Urine protein-creatinine index, ACEI: Angiotensin-converting enzyme inhibitor, ARB: Angiotensin receptor blockers, NA: Not available, MMF: Mycophenolate mofetil, PLEX: plasma exchange, IVIG: intravenous immunoglobulin *log copies/mcg total RNA

| No. | Urine<br>IP-10* | ISN/<br>RPS<br>class | Foot<br>process<br>effacement | Crescent | Serum<br>Creatinine<br>(mg/dL) | Serum<br>albumin<br>(g/dL) | Urinary<br>WBC<br>(cells/hpf) | Urinary<br>RBC<br>(cells/hpf) | UPCI<br>(g/g) | Anti-dsDNA<br>(IU) | C3<br>(mg/dL) | C4<br>(mg/dL) | ACEI/ARB | Previous<br>immunosuppression |
| --- | --- | --- | --- | --- | --- | --- | --- | --- | --- | --- | --- | --- | --- | --- |
| Histologically active LN with <u>positive</u> urine IP-10 <u>without</u> proteinuric flare |  |  |  |  |  |  |  |  |  |  |  |  |  |  |
| 1 | 2.58 | 3 | Focal | Absent | 0.36 | 3.8 | 20 | 5 | 0.5 | 800 | 30 | 3.44 | None | Prednisolone<br>15 mg/day |
| 2 | 3.42 | 4 | NA | Present | 1.24 | 3.1 | 20 | 5 | 0.59 | 800 | 42.3 | 6.6 | None | Prednisolone<br>2.5 mg/day |
| 3 | 3.60 | 4 | NA | Absent | 0.76 | 4.3 | 1 | 0 | 0.31 | NA | 80.8 | NA | Enalapril<br>20 mg/day | MMF 1000 mg/day<br>Prednisolone 5 mg/day |
| 4 | 3.97 | 4 | Focal | Present | 2.1 | 3.6 | 50 | 10 | 0.28 | NA | NA | NA | None | Methylprednisolone IV<br>500 mg<br>Prednisolone 30mg/day |
| Histologically active LN with <u>negative</u> urine IP-10 <u>with</u> proteinuric flare |  |  |  |  |  |  |  |  |  |  |  |  |  |  |
| 1 | 2.02 | 3+5 | Diffuse | Absent | 0.67 | 2.2 | 5 | 20 | 7.2 | 151.94 | 38.1 | 8.9 | Enalapril<br>10 mg/day | MMF 1000 mg/day<br>Prednisolone 60mg/day |
| 2 | 1.35 | 3 | Diffuse | Absent | 2.63 | NA | 20 | 100 | 1.62 | 800 | 56 | 10.4 | None | MMF 2000 mg/day<br>Prednisolone 50mg/day |
| 3 | 0.89 | 3 | NA | Absent | 3.62 | 3.7 | 2 | 5 | 9.77 | 0 | 39.6 | 4.84 | Candesartan<br>8 mg/day | PLEX, IVIG, rituximab,<br>methylprednisolone |

### The relationship between the level of urine IP-10 and the response to induction therapy

Of 36 patients with monthly urine IP-10 measurements for 6 months after being diagnosed with active LN, 29 (81%) responded to induction therapy at month 6. Urine IP-10 levels were similar between responders and non-responders at baseline (2.78 (95%CI 2.55-3.00) versus 2.90 (95% CI 2.13-3.67) log copies/mcg total RNA, p=0.7), and at month 6 (1.39 (95% CI 0.97-1.82) versus 1.69 (95%CI 0.97-2.41) log copies/mcg total RNA, p=0.4). The mean change of urine IP-10 level from baseline to month 6 was also comparable between responders and non-responders (−1.38 (95% CI −1.86 to −0.90) versus −1.21 (95% CI −1.99 to − 0.43) log copies/mcg total RNA, p=0.7).

### Positive urine IP-10 levels in patients with clinically inactive LN are associated with future LN flares

Of 46 patients with clinically inactive LN, 15 patients were followed longitudinally. LN flares occurred in 4 out of 8 patients with positive urine IP-10 (median follow-up time was 20.9 months (IQR 10.7 - 35.1) and median time to LN flare was 20.9 months (range 6.7 - 24.3)). In contrast, none of the seven clinically inactive patients with negative urine IP-10 developed LN flares (median follow-up time 19.4 months (IQR 17.5 - 43.7)).

## Discussion

The disparity between clinical presentation and pathological findings of LN has been well documented. Thus, novel biomarkers that accurately reflect histological activity in LN are much needed. In this study, we examined the utility of urine IP-10 in predicting histologic activity of LN, as well as its prognostic value for LN flare and treatment response. Clinicopathological discordance was found in 16% of biopsied patients. Urine IP-10 outperformed proteinuric flare in distinguishing histologically active LN (91% versus 84%), especially in those with clinicopathologic discrepancy. Positive urine IP-10 was also associated with higher risk of future LN flare in clinically inactive LN patients, compared to those with negative urine IP-10.

According to previous studies, approximately 25-50% of SLE patients had proliferative LN despite being clinically silent.(15, 16) Our study found that proteinuric flare was absent in 10% of patients with histologically active LN, where within this group of patients preserved podocyte integrity was observed in the cases that were inspected using electron microscope. On the other hand, 7% of biopsied patients had proteinuria from glomerulosclerosis. Urine IP-10 correctly predicted histological activity in the majority of these patients, which suggests the potential role of urine IP-10 in non-invasive monitoring of LN histologic activity.

Although the majority of clinically inactive LN patients in our study did not have a renal biopsy, a subset of these patients was followed longitudinally. Within 2 years, half of clinically inactive LN patients who had a positive baseline urine IP-10 developed LN flare, whereas none with negative urine IP-10 did. We hypothesize that some of the clinically inactive LN patients with positive urine IP-10 had smoldering LN that became overt after the immunosuppressive treatment was reduced. Consistent with our observations, a recent study found that patients in complete remission with residual histologic activity identified by repeat kidney biopsy experienced relapse after maintenance immunosuppression was tapered (17). In contrast, none of histologically active LN patients who had positive urine IP-10 without proteinuric flare developed clinical LN flare after long-term follow up. All of them received induction therapy despite being clinically inactive, which possibly prevented clinically overt LN. Therefore, urine IP-10 monitoring may be useful in identifying patients who are at risk of relapse and guiding immunosuppressive adjustments during the maintenance phase to avoid further LN flare and renal damage.

Nevertheless, urine IP-10 results should be interpreted with caution in patients receiving high dose immunosuppressive treatment, as we observed negative urine IP-10 in histologically active patients receiving prednisolone 50-60 mg/day. Because glucocorticoids are known to inhibit IFN-gamma signaling (18), IP-10 secretion in response to IFN-gamma may be suppressed. Our findings also do not support the use of urine IP-10 to predict treatment response, where the level of urine IP-10 at 6 months did not differ between responders and non-responders. However, it is important to note that the treatment response was defined by clinical criteria and may not accurately represent histological activity. A previous study found that histologically active LN persisted despite clinical response in nearly half of the patients, while one-third of clinical non-responders had inactive lesions on repeated biopsy. (19) Urine IP-10 may reflect histological activity, but this is difficult to prove without repeat renal biopsy.

The strengths of this study are having a relatively large number of patients, being able to perform renal biopsy in most patients with clinically active LN and some who were clinically inactive, having a standardized protocol for LN management and having a collection of longitudinal follow-up data. Limitations to our study include the following: Firstly, renal biopsies were not performed in most of the clinically inactive LN patients as well as after induction therapy. Secondly, patients with active non-renal SLE were not included in our sample, therefore, we cannot conclude that urine IP-10 is specific to active LN rather than the overall disease activity. Prior studies found no difference in urine IP-10 levels between active renal and nonrenal SLE (11, 14). However, these studies predominantly used clinical parameters to determine renal disease activity. Thus, it was difficult to discern whether high urine IP-10 levels were related to systemic activity of SLE or smoldering LN. Thirdly, there was no pure membranous LN in our study for comparison of urine IP-10 levels between proliferative and non-proliferative forms of LN. Similar to other studies, we previously reported that urine IP-10 was significantly lower in membranous LN compared to proliferative LN.(10, 20) Fourthly, we did not serially monitor urine IP-10 before the development of LN flare to evaluate any changes in urine IP-10 in relation to overt LN flare.

In summary, this study produced objective evidence that urine IP-10 may be a useful indicator of histological activity of LN. These findings may aid in the diagnosis of histologically active LN, particularly in patients without proteinuric flare or persistent proteinuria due to renal scarring. Urine IP-10 monitoring may also serve as a guide for immunosuppressive drug adjustment to prevent LN flare and eventually improve long-term renal outcomes. To improve the study’s validity, future research on urine IP-10 and other novel biomarkers should include surveillance or protocol-driven renal biopsy regardless of clinical disease activity.

## Data Availability

All data produced in the present study are available upon reasonable request to the authors

## Acknowledgements

This work was supported by the Thailand Research Fund (grant number MRG6180158); the National Science and Technology Development Agency and the Health Systems Research Institute (grant number FDA-CO-2562-9090-TH).

## Reference

1. Tektonidou MG, Dasgupta A, Ward MM. Risk of End-Stage Renal Disease in Patients With Lupus Nephritis, 1971–2015: A Systematic Review and Bayesian Meta-Analysis. Arthritis & rheumatology (Hoboken, NJ). 2016;68(6):1432–41.

2. Miranda-Hernandez D, Cruz-Reyes C, Angeles U, Jara LJ, Saavedra MA. Prognostic factors for treatment response in patients with lupus nephritis. Reumatologia clinica. 2014;10(3):164–9.

3. Rovin BH, Birmingham DJ, Nagaraja HN, Yu CY, Hebert LA. Biomarker discovery in human SLE nephritis. Bull NYU Hosp Jt Dis. 2007;65(3):187–93.

4. Moroni G, Radice A, Giammarresi G, Quaglini S, Gallelli B, Leoni A, et al. Are laboratory tests useful for monitoring the activity of lupus nephritis? A 6-year prospective study in a cohort of 228 patients with lupus nephritis. Ann Rheum Dis. 2009;68(2):234–7.

5. Zabaleta-Lanz M, Vargas-Arenas RE, Tapanes F, Daboin I, Atahualpa Pinto J, Bianco NE. Silent nephritis in systemic lupus erythematosus. Lupus. 2003;12(1):26–30.

6. Aten J, Roos A, Claessen N, Schilder-Tol EJM, Ten Berge IJM, Weening JJ. Strong and selective glomerular localization of CD134 ligand and TNF receptor-1 in proliferative lupus nephritis. J Am Soc Nephrol. 2000;11(8):1426–38.

7. Shiozawa F, Kasama T, Yajima N, Odai T, Isozaki T, Matsunawa M, et al. Enhanced expression of interferon-inducible protein 10 associated with Th1 profiles of chemokine receptor in autoimmune pulmonary inflammation of MRL/lpr mice. Arthritis Res Ther. 2004;6(1):R78–R86.

8. Wenzel J, Worenkamper E, Freutel S, Henze S, Haller O, Bieber T, et al. Enhanced type I interferon signalling promotes Th1-biased inflammation in cutaneous lupus erythematosus. J Pathol. 2005;205(4):435–42.

9. Hancock WW, Lu B, Gao W, Csizmadia V, Faia K, King JA, et al. Requirement of the chemokine receptor CXCR3 for acute allograft rejection. J Exp Med. 2000;192(10):1515–20.

10. Avihingsanon Y, Phumesin P, Benjachat T, Akkasilpa S, Kittikowit V, Praditpornsilpa K, et al. Measurement of urinary chemokine and growth factor messenger RNAs: a noninvasive monitoring in lupus nephritis. Kidney Int. 2006;69(4):747–53.

11. Abujam B, Cheekatla S, Aggarwal A. Urinary CXCL-10/IP-10 and MCP-1 as markers to assess activity of lupus nephritis. Lupus. 2013;22(6):614–23.

12. Narumi S, Takeuchi T, Kobayashi Y, Konishi K. Serum levels of ifn-inducible PROTEIN-10 relating to the activity of systemic lupus erythematosus. Cytokine. 2000;12(10):1561–5.

13. Puapatanakul P, Chansritrakul S, Susantitaphong P, et al. Interferon-Inducible Protein 10 and Disease Activity in Systemic Lupus Erythematosus and Lupus Nephritis: A Systematic Review and Meta-Analysis. Int J Mol Sci. 2019;20(19):4954.

14. El-Gohary A, Hegazy A, Abbas M, Kamel N, Nasef SI. Serum and Urinary Interferon-Gamma-Inducible Protein 10 in Lupus Nephritis. J Clin Lab Anal. 2016;30(6):1135–8.

15. Parikh SV, Almaani S, Brodsky S, Rovin BH. Update on Lupus Nephritis: Core Curriculum 2020. Am J Kidney Dis. 2020;76(2):265–81.

16. Shonrock AC, Rosa RG, Weigert S, Skare TL. Findings of renal biopsy in lupus patients with low levels of proteinuria. Acta Reumatol Port. 2010;35(3):399–400.

17. De Rosa M, Azzato F, Toblli JE, et al. A prospective observational cohort study highlights kidney biopsy findings of lupus nephritis patients in remission who flare following withdrawal of maintenance therapy. Kidney Int. 2018;94(4):788–794.

18. Hu X, Li WP, Meng C, Ivashkiv LB. Inhibition of IFN-gamma signaling by glucocorticoids. J Immunol. 2003;170(9):4833–9.

19. Zickert A, Sundelin B, Svenungsson E, Gunnarsson I. Role of early repeated renal biopsies in lupus nephritis. Lupus Sci Med. 2014;1(1):e000018.

20. Zeid MMH, Baddour NM, El-Neily DAE-M, Elshair HS, Mamdouh M. Study of urinary interferon gamma-induced protein 10 (IP-10) and urinary soluble CD 25 (sCD25) as markers of lupus nephritis and their relation to histological class. Alexandria Journal of Medicine. 2018;54(4):647–53

